# Exploration of the gut microbiome in Thai patients with major depressive disorder uncovered a specific bacterial profile with depletion of the *Ruminococcus* genus as a putative biomarker

**DOI:** 10.1101/2022.11.06.22282014

**Authors:** Michael Maes, Asara Vasupanrajit, Ketsupar Jirakran, Pavit Klomkliew, Prangwalai Chanchaem, Chavit Tunvirachaisakul, Sunchai Payungporn

## Abstract

Maes et al. (2008) published the first paper demonstrating that major depressive disorder (MDD) is accompanied by abnormalities in the microbiota-gut-brain axis, as evidenced by elevated serum IgM/IgA to lipopolysaccharides (LPS) of Gram-negative bacteria, such as *Morganella morganii* and *Klebsiella Pneumoniae*. The latter aberrations, which point to increased gut permeability (leaky gut), are linked to activated neuro-immune and oxidative pathways in MDD. To delineate the profile and composition of the gut microbiome in Thai patients with MDD, we examined fecal samples of 32 MDD patients and 37 controls using 16S rDNA sequencing and analyzed α-(Chao and Shannon indices) and β-diversity (Bray-Curtis dissimilarity) and conducted Linear discriminant analysis (LDA) Effect Size (LEfSe) analysis. Neither α-nor β-diversity differed significantly between MDD and controls. *Rhodospirillaceae, Hungatella, Clostridium bolteae, Hungatella hathewayi*, and *Clostridium propionicum* were significantly enriched in MDD, while Gracillibacteraceae family, *Lutispora, and Ruminococcus genus, Ruminococcus callidus, Desulfovibrio piger, Coprococcus comes*, and *Gemmiger*, were enriched in controls. Contradictory results have been reported for all these taxa, with the exception of *Ruminococcus* which is depleted in 6 different MDD studies (one study showed increased abundance), many medical disorders that show comorbidities with MDD, and animal MDD models. Our results may suggest a specific profile of compositional gut dysbiosis in Thai MDD patients with increases in some pathobionts and depletion of some beneficial microbiota. The results suggest that depletion of *Ruminococcus* may be a more universal biomarker of MDD that maybe contributes to increased enteral LPS load, LPS translocation, and gut-brain axis abnormalities.

## Introduction

There is now evidence that major depressive disorder (MDD) is a neuro-immune, neuro-oxidative and neuro-nitrosative (NINONS) disorder characterized by a) activation of the immune-inflammatory responses system (IRS) and a relative defcit in the compensatory immune-regulatory system (CIRS) and T regulatory (Treg) cell functions, which tend to attenuate an overzealous IRS; b) activation of oxidative pathways causing damage to lipids, proteins and DNA and IgM/IgG-mediated autoimmune responses to self-antigens and oxidative specific epitopes; c) hypernitrosylation with increased IgM-mediated autoimmune responses to nitrosylated proteins; and d) lowered antioxidant levels, including lowered high density lipoprotein and lowered lecithin acyl transferase and paraoxonase-1 (PON1) activity [1-8].

Despite the fact that MDD is a NINONS-associated disorder, the primary question is what causes these pathophysiological deviations [9]. Major contributors are psychosocial stressors, particularly adverse childhood experiences [8], genetic polymorphism, for example the PON1 Q192 gene variant [8], nutritional factors, including lowered omega-3 polyunsaturated fatty acids [4], zinc and vitamin D [10]; tobacco use disorder [9], metabolic aberations [11], and increased load of lipopolysaccharides (LPS) due to periodontitis [12] or translocation of Gram-negative gut-commensal bacteria via increased permeability of the intestinal barrier [13].

In fact, the first paper indicating that MDD is associated with alterations in the gut microbiota – brain axis was published in 2008, stating that many, but not all, MDD patients exhibit increased translocation of Gram-negative enterobacteria or their LPS as assessed with increased serum levels of IgM/IgA to the LPS of *Morganella morganii, Hafnia alvei, Citrobacter Koseri, Pseudomonas Putida, Pseudomonas Aeruginosa* and *Klebsiella Pneumoniae* [13]. In addition, we discovered that in numerous case reports, these elevated IgM/IgA responses to PLS were accompanied by indicators of gut dysbiosis, such as a dysbalance in the gut flora and changes in secretory IgA, β-defensin, α-antitrypsin, and calprotetine levels in stool [14,15]. The increased bacterial translocation in MDD is associated with greater severity of depression and irritable bowel symptoms, and is frequently associated with small intestine bacterial overgrowth (SIBO), as well as food, lactose, fructose, and gluten intolerances, according to case reports [14,15]. Importantly, in MDD, there are highly significant associations between IgA/IgM responses to LPS of gut commensal bacteria (indicating leaky gut) and NINONS and autoimmune pathways, indicating that bacterial translocation in MDD may drive, at least in part, the activated NINONS pathways [16]. The stimulation of the Toll-Like Receptor-4 (TLR4) complex by LPS, resulting in the activation of NINONS pathways, is one mechanism by which leaky gut may produce depressive behaviors [17]. Moreover, TLR4 gene polymorphisms are associated with MDD [18] and increased stress-induced bacterial translocation stimulates CNS neuro-inflammatory pathways in a rodent depression model [19].

Recent studies in MDD have supported the primary findings of Maes et al. [13,14,16]. For instance, in MDD, greater intestinal permeability as measured by the lactulose/mannitol test is substantially related to depression severity [20]. MDD is also characterized by other biomarkers of leaky gut, such as elevated LPS-binding protein and intestinal fatty acid-binding protein levels in association with inflammatory biomarkers and higher depression severity [21,22]. A study published in Science of how the diet modulates the microbiome, observed increased levels of *Morganella* and *Klebsiella* in association with mental depression [23]. MDD is related with leaky gut indicators in patients with inflammatory bowel disease [24], and, in MDD, bacterial translocation is accompanied by a reduction in the CIRS functions, namely the number of Treg cells, thereby contributing to inflammatory responses [25]. A recent meta-analysis shows that major neuropsychiatric disorders including a depressive episode, schizophrenia and chronic fatigue syndrome, are characterized by increased zonulin (4 studies), LPS (2 studies), antibodies to LPS (7 studies), sCD14 (six studies), and LPS-binding protein (two studies) [26].

Several etiologic factors can lead to leaky gut, including inflammatory processes, oxidative stress, nutritional factors, use of antibiotics and nonsteroid anti-inflammatory drugs, alcohol abuse, chemotherapy, surgery, radiation, viral infections including HIV, IBD, autoimmune disorders, and compositional gut dysbiosis with variations in gut microbiota communities [27]. Changes in gut microbial populations are now proposed to be strongly implicated in the pathophysiology of MDD [28,29]. Using second-generation sequencing of bacterial 16S RNA genes, for instance, it was discovered that the alpha diversity of gut bacteria was lower in MDD than in controls [30]. A recent review demonstrates that MDD is associated with a disparate representation of bacterial genus in comparison to controls, including increases in *Klebsiella, Clostridium, Blautia, Parabacteroides, Parasutterella, Streptococcus, Anaerostipes, Lachnospiraceaeincertaesedis*, and *Phascolarctobacterium*, and decreases in *Ruminococcus, Faecalibacterium, Bifidobacterium, Dialister*, and *Escherichia/Shigella* [31]. At the phylum level Actinobacteria, Bacteroidetes, Firmicutes, Fusobacteria, and Protobacteria were different between MDD and controls [31]. Linear Discriminant Analysis Effect Size (LEfSe) showed that Prevotellaceae and *Prevotella* showed increased abundance in MDD, whereas Bacteroidaceae, *Bacteroides*, and *uncultured_Mesorhizobium_sp*. showed increased abundance in controls [32].

Functional gut dysbiosis may further contribute to MDD via many different pathways. First, via increased abundance of pathobionts, which may contribute to increased NINONS activities, sympatho-adrenal-system activity, metabolic changes, insulin resistance, neurodegenerative processes, and damage to lipoproteins. Second, by depletion of anti-inflammatory microbiota and microbiota that keep the epithelial barrier and the gut healthy. These include microbiota that produce alkaline phosphatase, microbiota that break down polysaccharides and LPS, and sulphate, and other beneficial microbiota that generate short-chain fatty acids (SCFAs, like butyrate) and vitamin antioxidants [33-53]

Nevertheless, there is no data whether in Thai MDD patients there are any indicants of gut dysbiosis, as indicated by reduced diversity of gut flora, and differential abundance of bacterial taxa. Hence, the present study was performed to delineate whether MDD is characterized by diminished gut bacterial microbiota alpha and beta diversity, changes in relative abundances of gut bacteria at the phylum, genus, and species levels, and differential abundance of bacterial taxa based on LEfSe.

## Materials and Methods

### Participants

For this study, thirty-seven normal controls and thirty-two patients with major depressive disorder (MDD) were recruited from the Department of Psychiatry’s outpatient clinic at King Chulalongkorn Memorial Hospital in Bangkok, Thailand. Participants ranged in age from 19 to 58 years old and were of both sexes. They were diagnosed with MDD using DSM-5 criteria. The control group was recruited in the same catchment area as the patients, Bangkok, Thailand, via word of mouth. Control participants having any DSM-5 axis 1 disorder diagnosis or a positive family history of MDD, bipolar disorder (BD) or suicide were excluded from the study. MDD participants having any DSM-5 axis 1 disorder diagnosis other than MDD were excluded from the study, e.g. BD, schizophrenia, schizo-affective disorder, anxiety disorders, obsessive compulsive disorder, post-traumatic stress disorder, autism, substance use disorder (except nicotine dependence), and psycho-organic disorders. Patients and controls were excluded if they had any of the following conditions: a) (auto)immune diseases, such as chronic obstructive pulmonary disease, cancer, psoriasis, type 1 diabetes, and asthma, b) inflammatory bowel disease or irritable bowel syndrome; c) neurodegenerative, neuroinflammatory, or neurological disorders, such as Alzheimer’s disease, multiple sclerosis, stroke, epilepsy, or Parkinson’s disease; d) inflammatory or allergic reactions three months prior to the study; e) pregnant or lactating women.

All patients and controls provided written consent before taking part in this study. The study was carried out in compliance with international and Thai ethical standards as well as privacy legislation. The Chulalongkorn University Faculty of Medicine’s Institutional Review Board in Bangkok, Thailand (#446/63) approved the study in accordance with the International Guidelines for the Protection of Human Subjects as required by the Declaration of Helsinki, The Belmont Report, the CIOMS Guideline, and the International Conference on Harmonization in Good Clinical Practice (ICH-GCP).

### Clinical assessments

Semi-structured interviews were conducted by a trained research psychologist specialized in mood disorder research. To assess the severity of depression symptoms, we employed the Hamilton Depression Rating Scale (HDRS) [53] and the Beck Depression Inventory-II (BDI) [54]. To assess the axis-1 diagnosis, the Mini-International Neuropsychiatric Interview (M.I.N.I.) was utilized [55]. Using DSM-5 criteria, tobacco use disorder (TUD) was diagnosed. Body mass index (BMI) was calculated by dividing body weight (in kilograms) by length squared (in meter).

### Assays

#### Fecal sample collection and DNA extraction

Approximately 20 mg of fecal samples were collected in sterile test tubes containing 2 mL of DNA/RNA Shield™ reagent (ZYMO Research, USA) and kept at -20°C until tested.

DNA was extracted using ZymoBIOMICS™ DNA Miniprep Kit (ZYMO Research, USA) following the manufacturer’s standard protocol.

#### 16S rDNA amplification

The full length of the bacterial 16S rDNA gene (1.5 kb) was amplified by PCR using specific primers: 5′-TTTCTGTTGGTGCTGATATTGCAGRGTTYGATYMTGGCTCAG-3′ and ′-ACTTGCCTGTCGCTCTATCTTCCGGYTACCTTGTTACGACTT-3′ as described previously [56]. The first round of PCR reaction contained 1 µg of DNA template, 0.2 µM of each primer, 0.2 mM of dNTPs, 1X Phusion™ Plus buffer, 0.4 U of Phusion Plus DNA Polymerase (Thermo Scientific, USA) and nuclease-free water in a final volume of 20 µL. The PCR reaction was performed under the following thermal conditions: 98°C for 30 s; 25 cycles of amplification (98°C for 10 s, 60°C for 25 s, 72°C for 45 s) and followed by 72°C for 5 min. After that, the barcodes were attached to the 16S rDNA amplicon by 5 cycles of amplification (98°C for 10 s, 60°C for 25 s, 72°C for 45 s) based on PCR Barcoding Expansion 1–96 (EXP-PBC096) kit (Oxford Nanopore Technologies, UK). The amplicons were purified using QIAquick^®^ PCR Purification Kit (QIAGEN, Germany) according to the manufacturer’s protocol.

#### 16S rDNA amplicon sequencing based on Oxford Nanopore Technology (ONT)

The concentrations of purified amplicons were measured using a Qubit 4 fluorometer with Qubit dsDNA HS Assay Kit (Thermo Scientific, USA). Then the amplicons with different barcodes were pooled at equal concentrations and purified using 0.5X Agencourt AMPure XP beads (Beckman Coulter, USA). After that, the purified DNA library was end-repaired and adaptor-ligated using Ligation Sequencing Kit (SQK-LSK112) (Oxford Nanopore Technologies, UK). Finally, the library was sequenced by the MinION Mk1C platform with R10.4 flow cell (Oxford Nanopore Technologies, UK).

### Data processing and statistical analysis

Guppy basecaller software v6.0.7 [57] (Oxford Nanopore Technologies, UK) was used for base-calling with a super-accuracy model to generate pass reads (FASTQ format) with a minimum acceptable quality score (Q > 10). The quality of reads was examined by MinIONQC [58]. Then, FASTQ sequences were demultiplexed and adaptor-trimmed using Porechop v0.2.4 [59]. The filtered reads were then clustered, polished, and taxonomically classified by NanoCLUST [60] based on the full-length 16S rRNA gene sequences from the Ribosomal Database Project (RDP) database [61]. The abundance taxonomic assignment data were converted into QIIME data format to illustrate bacterial richness and evenness based on their taxa abundances and alpha diversity (Chao1 and Shannon indexes). Then, the beta diversity was analyzed with the Bray-Curtis cluster analysis index using a plug-in implemented for QIIME2 software v2021.2 [62]. The normalized data were visualized by MicrobiomeAnalyst [63]. The differential abundance was analyzed based on the Linear discriminant analysis Effect Size (LEfSe) with *P* < 0.05 and Linear discriminant analysis (LDA) score > 2 [64] by using the Galaxy server [65].

## Results

### Socio-demographic and clinical data

**Table 1** displays the sociodemoghraphic data of the participants divided into controls and MDD patients. There were no differences in age, sex ratio, education, BMI, MetS, and employment among both groups. There was a trend to a lower rate of married people, and increased TUD rate in MDD patients as compared with controls. The HDRS and BDI scores were significantly higher in MDD patients than in controls. Part of the MDD patients were treateted with psychotropic drugs, namely sertraline (n=10), fluoxetine (n=6), escitalopram (n=6), trazodone (n=4), benzodiazepines (n=11), antipsychotic agents (n=5) and mood stabilizers (n=1). Statistical analyses were used to look if the drug state of the patients might affect the microbiome features. However, even without false discovery rate p correction no significant effects of these psychotropic drugs could be found on the transformed (insometric log ratio trasnsformation using Box-Cox transformation) microbiota data (at the phylum, genus and species level).

**Table 1.**
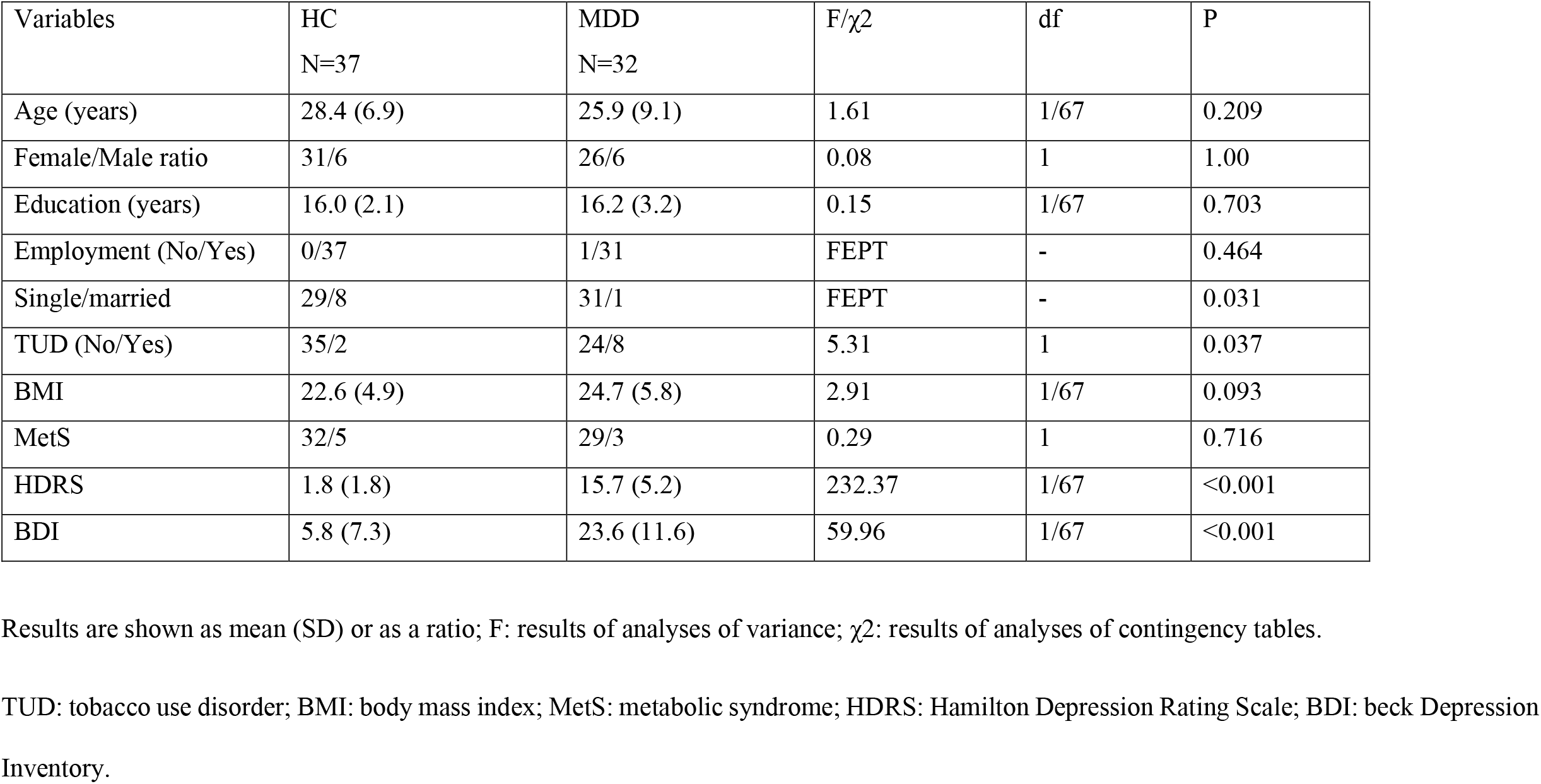
Socio-demographic and clinical data of patients with major depressive disorder (MDD) and healthy controls (HC) included in the present study.

### Gut bacterial microbiota diversity

The full-length bacterial 16S rDNA was sequenced based on high throughput long-read nanopore sequencing, providing 827,392 raw reads in total, with an average of 13,133 reads per sample. After quality filtering, the retained reads were classified into operational taxonomic units (OTUs). Alpha diversities (Chao1 and Shannon indexes) were used to demonstrate the richness and evenness of bacterial comparisons between controls and MDD groups based on their relative abundances. There were no significant differences using the the Mann–Whitney U test between both study groups, as shown in **Figure 1A-B**. Based on a Bray–Curtis dissimilarity index to compare the bacterial communities between controls and MDD groups (Figure 1C), the result showed no significant differentiation with identity at a 95% confidence interval.

**Figure 1.**
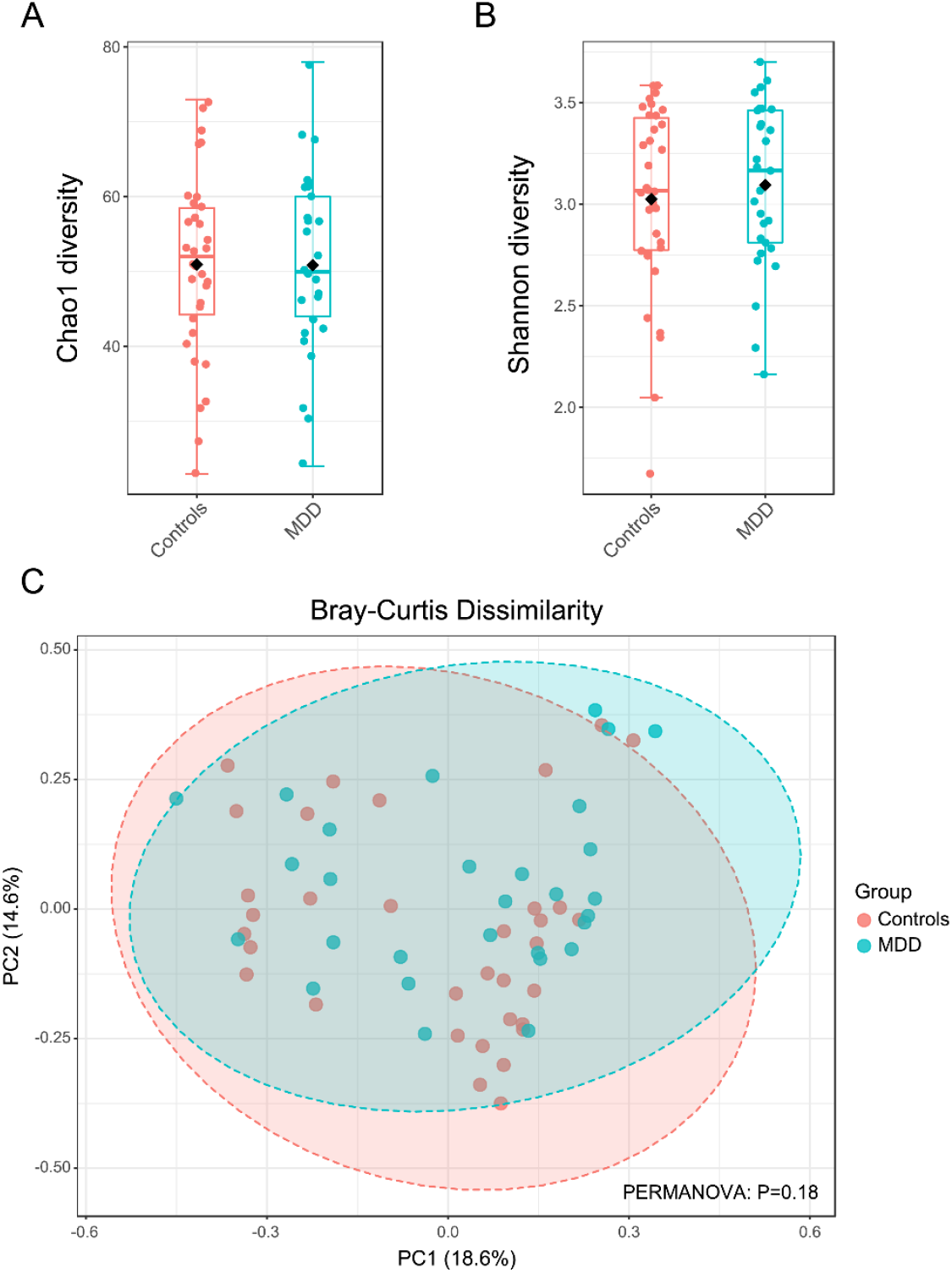
Gut bacterial microbiota diversity in controls (pink) and MDD (blue) groups. The alpha diversity comparison calculated by Chao1 (A) and Shannon (B) indexes were shown as box plots with the error bars representing the standard deviation and calculated statistically significant difference by Mann–Whitney U test. The beta diversity was presented by principal coordinate analysis (PCoA) plots based on Bray-Curtis distance (C). Permutational ANOVA (PERMANOVA) was applied to calculate statistical differences in beta diversity.

### Gut bacterial profile and composition

The gut bacterial profiles were characterized by the microbial composition of samples in the controls and MDD groups. **Figure 2** shows that the most abundant phylum was Firmicutes, followed by Bacteroidetes and Proteobacteria in both groups. The top three most abundant bacteria at the genus level were *Faecalibacterium, Prevotella* and *Bacteroides* in the controls (68.9%) and the MDD group (54.7%). Additionally, relative abundance in species level between groups could not be clearly distinguished. However, *Faecalibacterium prausnitzii, Prevotella copri, Oscillibacter valericigenes, Bacteroides vulgatus* and *Prevotella stercorea* were the most prevalent gut bacteria in the controls (61.3%) and the MDD group (41.9%).

**Figure 2.**
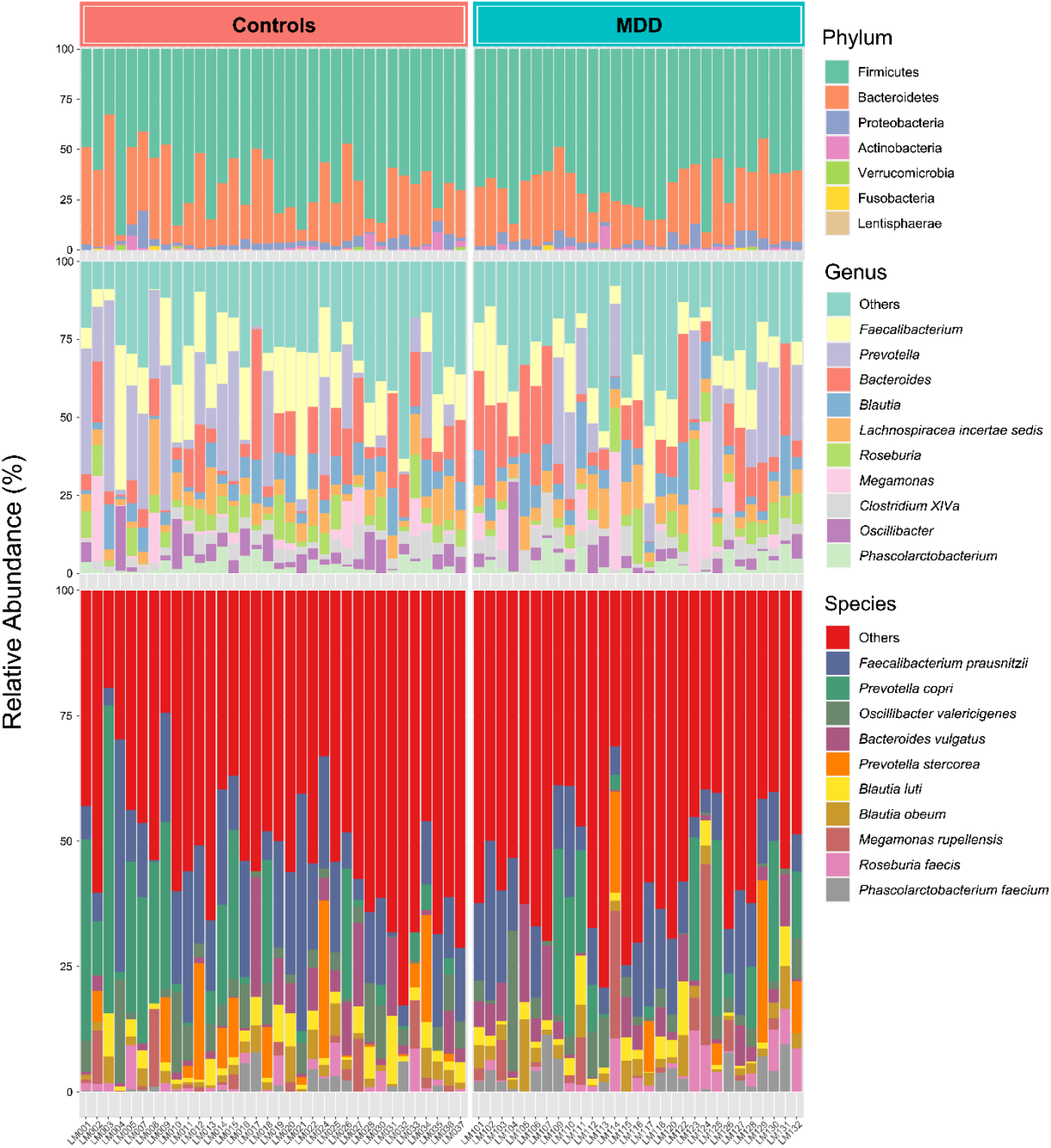
The stacked plot showed relative abundances of gut bacteria at the phylum, genus and species levels between controls and MDD groups. The data were normalized by a total sum scaling (TSS) method. The colored bars represent various bacterial taxa identified in the gut bacterial profiles.

### Differential abundance of gut bacteria between controls and MDD groups

LEfSe analysis was performed to classify the significant differences in bacteria between controls and MDD groups based on Linear discriminant analysis (LDA) scores (> 2), as shown in the bar graph (**Figure 3A**) and cladogram (Figure 3B). The Rhodospirillaceae family, *Hungatella* genus, and bacterial species, including *Clostridium bolteae, Hungatella hathewayi* and *Clostridium propionicum*, were significantly enriched in the MDD group. *Desulfovibrio piger, Ruminococcus callidus, Coprococcus comes, Gemmiger formicilis* and *Phascolarctobacterium succinatutens* were enriched in the control group.

**Figure 3.**
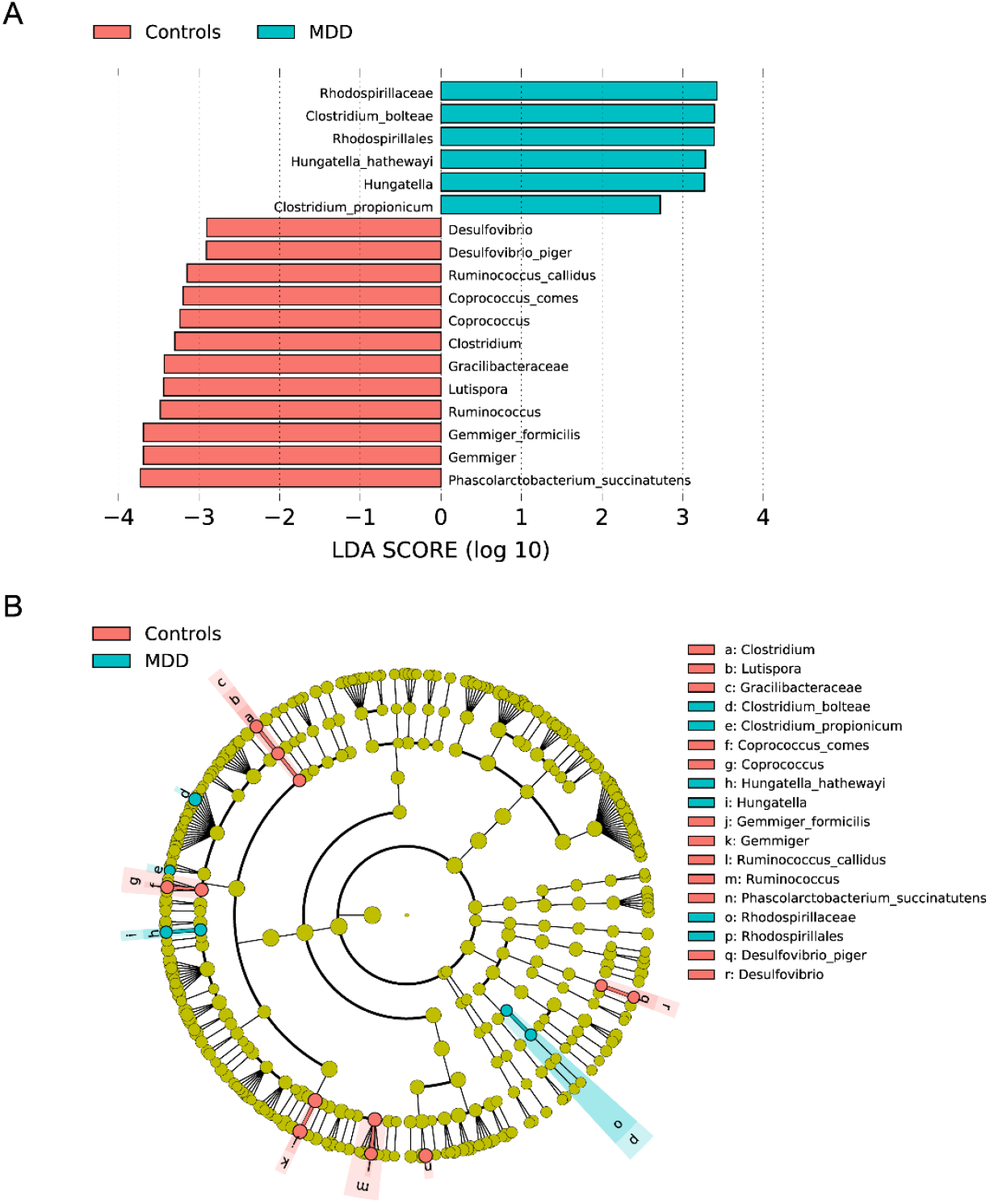
The differential abundance of bacterial taxa between controls and MDD groups based on Linear discriminant analysis Effect Size (LEfSe). LDA scores significantly enriched in the differentially taxonomic levels among groups were represented as the bar graph (A) and Cladogram (B).

## Discussion

### Key findings of our study

The major findings of this study are that a) there are no significant differences in α-or β-diversity between MDD and controls; b) there is no clear difference in relative abundance of species between MDD and controls; and c) LEfSe analysis revealed that the Rhodospirillaceae family (Gram-negative, rod-shaped to spirillium-formed, purple non-sulfur bacteria which comprise 34 genera; [37,38]), *Hungatella* genus (anaerobic, Gram-postive bacterial genus from the family of *Clostridiaceae*; [39]), and the species *Clostridium bolteae* (anaerobic, Gram-positive, rod-shaped, spore-forming bacteria, [40]), *Hungatella hathewayi* (anerobic, Gram-postive bacterium, [41]), and *Clostridium propionicum* (anaerobic, Gram-positive, rod-shaped bacteria, [42]) were significantly enriched in the MDD group. In contrast, *Desulfovibrio piger (aerotolerant, Gram-negative bacteria*, [43]), *Ruminococcus callidus* (anaerobic, Gram-positive bacteria; [48]), *Coprococcus comes* (anaerobic Gram-postive, cocci, [44]), *Gemmiger formicilis* (anaerobic weakly Gram-postive bacteria; [66]), *Phascolarctobacterium succinatutens* (strictly anaerobic, Gram-negative bacteria; [46]), *Lutispora* (anaerobic, Gram-stain-negative and Gram-positive cell-wand structure; [67]) and Gracillibacteraceae (belong to the Furmicutes phylum) are enriched in the control group.

### α-diversity and MDD

Our negative findings regarding α-diversity corroborate some prior research papers. Four reviews on α-diversity in MDD or mixed groups of the major psychiatric diseases found no convincing evidence of changes in α-diversity of bacteria in MDD [68-71]. According to the most recent systematic review, there was a greater number of studies that found no differences (n = 8) or inconsistent results (n = 7) than those that revealed reduced α-diversity (n = 5). In MDD, three research papers found no differences, four studies observed inconsistencies, and two studies found decreased α-diversity [68]. According to a recent systematic review [72], 14 out of 21 studies identified no significant variations in α-diversity between MDD and controls. McGuinness et al. [71] stated that there is no convincing evidence that patients with mental illnesses show less α-diversity. Ritchie et al. [73] and Thapa et al. [74] reported that there are no significant differences in the α-diversity of bacteria between patients with MDD and controls, whereas other studies reported reduced [75] or increased α-diversity [76] in MDD. Ye et al. [77] determined, using the Chao1 and Shannon indices, that α-diversity was greater in MDD than in controls. In their investigation, Zhang et al. [32] discovered, using the Simpson and Pielou’s index, that the α-diversity was reduced in MDD patients without adverse childhood experiences, but no differences were found between controls and MDD patients with adverse childhood experiences. Overall, the results imply that the microbiome α-diversity is not significantly altered in MDD.

### β-diversity and MDD

Our negative findings regarding β-diversity corroborate previous studies. The recent systematic review of Borkent et al. (2022) showed that three research papers did not identify differences in β-diversity between MDD and controls, while four investigations reported decreased β-diversity [68]. Inconsistent β-diversity findings were also presented in the review by Simpson et al. [69]. In their systematic review, Alli et al. [72] found that 12 out of 18 studies demonstrated that β-diversity was significantly different between MDD and controls. Moreover, McGuinness et al. [71] delineated that studies on β-diversity in mental diseases, including MDD, are reasonably consistent. Some more recent studies have revealed contradictory findings regarding β-diversity in MDD. Thus, Ritchie et al. [73] and Thapa et al. [74] were unable to discover significant changes in β-diversity in MDD and adolescent depression versus controls, respectively. Liu et al. [75] observed a reduction in both α- and β-diversity of the gut microbiota composition. Sun et al. [76] discovered variations in β-diversity between MDD and controls and found that, at p=0.03, the between-group differences were greater than the within-group differences. Zhang et al. [32] in their β-diversity study, found significant differences between controls and MDD. Overall, our findings add to the negative research findings on β-diversity in MDD which account for roughly one-third of all studies.

### Abundance of gut microbiome taxa in MDD

Borkent et al. [68] reported that all research on MDD and other severe psychiatric diseases, such as schizophrenia and BD, reported statistically significant variations in the relative abundance of bacterial taxa. Nonetheless, it appears that these distinctions are sometimes extremely inconsistent, whilst the most consistent results were increased abundance of *Lactobacillus, Streptococcus*, and *Eggerthella* in controls, and decreased abundance of anti-inflammatory butyrate-producing *Faecalibacterium* in the combined group of mental disoders. Our assessment of the signature of gut microbiota utilizing LEfSe analysis to find the classes of organisms that explain differences across diagnostic groups did not support the conclusions of Borkent et al. [68]. Our review shows that the microbiome LEfSe profile of MDD has only a few parallels with previous LefSe investigations in MDD.

For instance, Zhang et al. [38] revealed that 27 microorganisms were linked to MDD or controls. These authors found that Ruminococcaceae family, *Gemmiger*, and *Phascolarctobacterium* were enriched in controls, which partly corresponds with our results showing that *Gemmiger, Gemmiger f*., and *Phascolarctobacterium s*. were enriched in controls. On the other hand, neither study discovered any similarities in the taxa related to MDD. In another study, multi-omics studies of the gut microbiome in MDD and LEfSe analysis identified numerous microbiome characteristics in MDD, however none of these correlated with our findings [78]. In addition, whereas we observed an increase in the prevalence of *Ruminococcus* and *Clostridium* in the control group, Zhao et al. [78] observed an increase in the prevalence of some *Ruminococcus* and *Clostridium* taxa in MDD. The LEfSe analysis in the study by Liu et al. [75] revealed that Deinococcaceae were enriched in MDD, whereas Clostridiaceae, Bacteroidaceae, Turicibacteraceae, and Barnesiellaceae were enriched in controls. At the genus level, *Deinococcus* and *Odoribacter* were associated with MDD, and *Clostridium, Bacteroides, Alistipes, Turicibacter, Roseburia*, and *Enterobacter* with controls. Thus, the sole resemblance between Liu’s study and ours is the higher relative abundance of *Clostridium* in the control group. We and Jiang et al. [79] both observed increased abundance in *Ruminococcus* in MDD patients.

LEfSe analysis performed on patients with depression and anxiety in the screening for gastro-intestinal cancer confirmed that *Gemmiger* and *Ruminococcus* were overabundant in controls, but none of the other taxa were discriminant features [80]. In a study examining the microbiome in a major depressive episode associated with BD, LEfSe analysis revealed differentially abundant features, including increased abundance of the phylum Actinobacteria and the class Coriobacteria, whereas *Faecalibacterium* genus and Ruminococcaceae family were more abundant in controls [81]. These latter findings corroborate our results that the *Ruminococcaceae* family is overexpressed in controls. Another study discovered fecal microbiome characteristics that differentiate childhood depression [82]. Although this LefSe analysis discovered numerous discriminative taxa at an LDA threshold of > 3, there were no parallels between our findings and those of Ling et al. Moreover, whilst we discovered that *Gemmiger* and *Desulfovibrio* were abundant in controls, Ling et al. [82] discovered the opposite. There are no similarities between our results (obtained in younger MDD patients and controls) and those of patients with late-life depression [83], despite the fact that the latter LEfSe analysis found many significant abundances of bacterial taxa at the genus level in controls as opposed to patients with late-life depression.

Overall, a comparison of the LEfSe analysis undertaken in MDD reveals few parallels between the various investigations. As the microbiome compositium is heavily impacted by the diet [84], it is likely that microbiome features of a human disorder are culture-specific and may not always correspond with the profiles established in other countries. Therefore, the micriobiome profile shown here may be unique to Thai individuals with MDD, except maybe for the *Ruminococcus* genus which has been detected in other countries.

### Is there compositional gut dysbiosis in MDD?

Certainly, it is plausible to hypothesize that the dysbiotic microbiome features identified by our LEfSe analysis contribute to the pathophysiology of MDD by a) increasing the predominance of some potential pathobionts, such as Rhodospirillaceae (inhibit calpain protease which play a role in neurodegenerative processes*), C. propionicum* (may induce the sympatho-adrenal-system with increased norepinephrine production and consequent insulin resistance), and *H. hathewayi* (possible pathogen that is associated with colorectal cancer); and b) lowering the abundance of protective microbiota including *Clostridium* (promote SCFA production), *Coprococcus* (involved in the maintenance of a healthy gut and epithelial barrier function), *Desulfovibrio* (reduce sulphate production which may be related to many physio-somatic symptoms when upregulated), *Phascolarctobacterium succinatutens* (production of SCFA including proprionate), Gracillibacteraceae (association with attention deficit disorder) and *Gemmiger* and *G. formicillis* (production of SCFA; depletion is associated with inflammatory bowel disease) [37-47, 66].

These data may imply compositional gut dysbiosis in MDD, which is defined as an increase in potentially harmful enterobacteria and a decrease in gut beneficial microbiota. However, it remains unknown if these changes in pathobionts and beneficial microbiota reflect functional gut dysbiosis and whether dysbiosis is a cause or effect of the NINONS-responses in MDD. If these results are indicative of functional dysbiosis, they are likely unique to Thai patients with MDD. In fact, distinct microbiome profiles in distinct cultures may result in comparable functional effects, either pathological or beneficial.

### Is depletion of Ruminococcus important to MDD?

Maybe the most consistent finding in the reviewed literature, is the higher relative abundance of *Ruminococcus*, which is increased in ours and some other studies [38,79, 80, 81], whereas one study reported contradictory results [78]. Additionally, utilizing bidirectional Mendelian randomization, Chen et al. [85] discovered that Ruminococcus protects against MDD. The chronic restraint mouse model of depression is characterized by the depletion of *Ruminoccocus*, gut dysbiosis, and leaky gut in conjunction with activated NINONS pathways and decreased neurotrophic protection in the brain [86]. In addition, treatment with crocin-I decreased depressive-like behaviors and these effects are mediated by increased abundance of beneficial microbiota, including *Ruminococcus*, improvement of leaky gut (effects on claudin-1 and occludin), and attenuation of activated NINONS pathways in the brain [86]. Interestingly, the abundance of *Ruminococcus* is significantly associated with 5-HT levels in a rodent model of postpartum depression [87]. Different *Ruminococcus* species are associated with NINONS-associated human disorders that exhibit a strong comorbidity with MDD [88], including chronic fatigue syndrome, inflammatory bowel disease, type 1 and 2 diabetes mellitus, atherosclerosis and cardiovascular disease, and nonalcoholic fatty liver disease (NAFLD) [47]. In Parkinson’s disease, those with cognitive impairment had a lower abundance of the genus *Ruminococcus* than those without cognitive abnormalities [89]. *Ruminococcus* genus and *R. callidus* are much less prevalent in the feces of patients with inflammatory bowel disease [49]. The *Ruminoccocus* genus is a key symbiont of the core gut microbiota and plays an important role in gut health via degradation and conversion of polysaccharides (e.g. cellulose and starch) into nutrients, which provide cross-feeding to other microbiota [48,49]. In addition, species of this genus display cellulolytic activity and may degrade LPS. Furthermore, different *Ruminococcus* species produce alkaline phosphatase [35] which improves gut barrier functions and detoxifies LPS thereby preventing activation of NINONS pathways [50,51]. One could, therefore, hypothesize that depletion of this genus may have contributed to lowered LPS breakdown and thus increased translocation of LPS in MDD particularly when deficits in tight junctions are present [13-16]. *R. callidus* is consistently present in the healthy human gut and probably plays a role in the maintenance of a healthy gut environment [49]. As such, depletion of *Reminococcus* could maybe constitute a more universal feature of MDD.

### Limitations

It would be fascinating to assess the functional repercussions of microbiome changes in MDD using shotgun metagenomic sequencing in conjunction with pathway enrichment analysis. Importantly, future research should employ Kegg pathway analysis to further delineate gut LPS biosynthesis in MDD as a risk factor contributing to the increased translocation of Gram-negative bacteria. Moreover, future prospective studies should explore the microbiome before and after the development of the first depressive episode in young adults at risk for developing depression (e.g., those with greater childhood adverse experiences and increased neuroticism scores).

## Conclusions

The levels of α- and β-diversity were similar between those with MDD and controls. The LEfSe analysis revealed an increased abundance of some pathobionts and a decrease in some beneficial gut commensals. Although these results may theoretically suggest compositional gut dysbiosis in MDD, previous studies have been unable to delineate the same microbiome features and some have even shown contradictory results. If our results would be replicated in other Thai samples, the LEfSe profile established here is likely to be unique to Thai patients with MDD. The depletion of *Ruminococcus* has been observed in some human studies (although one study reported contradictory results), animal models and disorders that are comorbid with MDD and, therefore, the most important takeaway from this study is that low levels of *Ruminococcus* may have a role in MDD. *Ruminococcus* and the species *R. callidus* serve a useful role in the maintenance of a healthy gut environment: they degrade polysaccharides into nutrients and degrade LPS, functions that could theoretically increase LPS translocation in the presence of preexisting deficiencies in the tight and adherens junctions of the paracellular pathway. In our opinion, the latter aberrations are secondary to the activation of NINONS-pathways in MDD, but depletion of beneficial gut commensal genera and species, including *Ruminococcus* and the species *R. callidus*, may contribute to increased LPS or bacterial translocation.

## Data Availability

The dataset generated during and/or analyzed during the current study will be available from MM upon reasonable request and once the authors have fully exploited the dataset.

## Author Declarations

### Conflicts of Interest

The authors declare that they have no known competing financial interests or personal relationships that could have influenced the work reported.

### Funding

This research was supported by a Rachadabhisek Research Grant, Faculty of Medicine, Chulalongkorn University, Bangkok, Thailand, to M.M. The sponsor had no role in the data or manuscript preparation.

### Author’s Contributions

All authors contributed to the paper. MM designed the study. Patients were recruited by AV, KJ and CT. Microbiome assays were performed by PK, PC and SP. Statistical analyses were performed by PK, SP and M.M. All authors revised and approved the final draft.

## Compliance with Ethical Standards

### Research involving Human Participants and/or Animals

This study was approved by the Institutional Review Board (IRB) of Chulalongkorn University, Bangkok, Thailand (IRB no. 446/63), which complies with the International Guideline for Human Research Protection as required by the Declaration of Helsinki.

### Informed consent

Before taking part in the study, all participants and/or their caregivers provided written informed consent.

## Notes

### Competing Interest Statement

The authors have declared no competing interest.

### Author Declarations

This study was approved by the Institutional Review Board (IRB) of Chulalongkorn University, Bangkok, Thailand (IRB no. 446/63), which complies with the International Guideline for Human Research Protection as required by the Declaration of Helsinki. Before taking part in the study, all participants and/or their caregivers provided written informed consent.

